# Olfactory training and visual stimulation assisted by web-application in patients with persistent olfactory dysfunction after SARS-CoV-2 infection: observational study

**DOI:** 10.1101/2021.05.13.21257176

**Authors:** Fabrice Denis, Anne-Lise Septans, Léa Périers, Jean-Michel Maillard, Florian Legoff, Hirac Gurden, Sylvain Morinière

## Abstract

**Background:** Persistent olfactory dysfunction (OD) is a significant complication of SARS-CoV-2 infection. Olfactory training (OT) using aromatic oils are recommended to improve olfactory recovery, but quantitative data are missing.

**Objective:** We aimed to quantify the benefit of OT associated with visual stimulation assisted by a dedicated web-application on patients with 1-month or more OD.

**Methods:** We performed an observational real-life data-based study on a cohort of patients with at least 1-month persistent OD included between 1/30/21 and 3/26/2021. Analysis was performed after a 4-weeks mean time of OT and at least 500 patients assessable for primary outcome. Participants exposed themselves twice daily to odors from four high concentration oils and visual stimulation assisted by a dedicated web-application. Improvement was defined as a 2/10 points increase on self-assessed olfactory visual analogue scale.

**Results:** 548 were assessable for primary outcome assessment. The mean baseline self-assessed olfactory score was 1.9/10 (SD 1.7) and increase to 4.6 (SD 2.8) beyond a mean time of olfactory training of 27.7 days (SD 17.2). Olfactory training was associated with at least 2-points increase in 64.2% (n=352). The rate of patients with improvement was higher in patients having trained for more than 28 days versus patients having trained for less than 28 days (72.2% vs 59.0% respectively, p=.002). The kinetic of improvement was 8 days faster in hyposmic than in anosmic patients (p<.001). The benefit was observed regardless of the duration of the OD.

**Conclusions:** OT associated with visual stimulation assisted by a dedicated web-application was associated with significant improvement in olfaction, especially if OT duration was superior to 28 days.

## INTRODUCTION

Anosmia is a frequent symptom of SARS-CoV-2 infection and its duration is usually less than two weeks before recovery. [1-3) However, at least 10% of patients will have persistent and chronic olfactory dysfunction such as diminished smell (hyposmia) or loss of smell (anosmia) which was shown to lead to decrease quality of life, depressive symptoms and nutrition issues. [4-6) One treatment option which is recommended against persistent olfactory dysfunction is the daily olfactory training using high concentration aromatic oils. [7] It showed significant results in postinfectious olfactory loss in a randomized controlled multicenter study. [8] In this trial, after 18 weeks olfactory training, function improved in 63% of patients having a duration of olfactory dysfunction of less than 12 months olfactory dysfunction using high concentration oils versus 19% in the control group using low concentration oils. Moreover, combination of visual stimulations to olfactory training may improve recovery results. [9]

No data about olfactory training in persistent olfactory dysfunction are available in SARS-CoV-2 patients with persistent olfactory dysfunction, but most of patients having 30-days or more hyposmia or anosmia seems to have a low rate of spontaneous recovery. [4]

In order to quantitatively study the time course of olfactory scores during olfactory training in real-life, we developed a web-application dedicated to olfactory training and visual stimulations as well as self-assessment and follow-up of olfactory scores. We assessed results in a real-life observational study.

## METHODS

The users were recruited via national media campaign in France including social media, radio and magazines between 1/30/2021 and 2/15/2021.

This observational data-based study was approved by the French National Health Data Institute which reviews ethical conduct of human subject’s research, data confidentiality, and safety. To participate, individuals were required to connect to the free covidanosmia.eu web-application and give electronic agreement. Respondents self-entered anonymously sociodemographic data, RT-PCR test results and diagnosis of SARS-CoV-2 olfactory dysfunctions. Patients were also asked to complete items about their co-morbidities, duration of olfactory symptoms and self-assessed intensity of olfactory dysfunction using subjective ratings with a visual analogue scale (0 no smell) to 10 (no smell alteration). [10] Patients were retained in the study analysis if they had a SARS-CoV-2 olfactory dysfunction, persistent from at least 1 month, with a reporting of at least 7 days of olfactory training and if their last olfactory function assessment on web-application diary was available. Exclusion criteria were normosmia (visual scale score >7/10), other causes of olfactory dysfunction such as chronic rhinosinusitis, nasal polyposis, allergic or idiopathic rhinitis, post-traumatic olfactory loss, or other acute or chronic nasal diseases (e.g., acute viral infections), malignant tumors or/and oncology therapies (radiation, chemotherapy), and history of surgery on the nose or paranasal sinuses.

Then, patients had to obtain the olfactory training kit from the application or from their pharmacist. Olfactory training was performed over a maximum period of 16 weeks. The web-application provides videos, tutorials for the training as well as periodic encouragements. Participants exposed themselves twice daily to odors from four high concentration oils: phenyl ethyl alcohol: rose odor from geranium rosa, eucalyptol: eucalyptus odor, citronellal: lemon odor, and eugenol: cloves odor. These four odorants were chosen to represent primary odor categories claimed by Henning [11,12]. They should sniff each odor for approximately 15 seconds blind and repeat this once with the name and a picture of the oil component on the screen by the application 30 seconds later (for example a picture of a lemon during lemon oil sniff)… Patients were asked to train in the morning and in the evening, resulting in a total of four expositions per day per odor. They were asked to keep a daily diary on the application where they rated overall olfactory abilities for each oil by subjective ratings with visual analogue scale.

We assessed the rate of self-assessed improvement of overall olfactory function along training time from data collected anonymously by web-application diary of patients. Improvement was defined as a 2/10 points or more increase on olfactory visual analogue scale. Study analysis was performed when mean time of olfactory training of the population was at least 4-weeks and when at least 500 patients were assessable for primary outcome.

Categorical variables were summarized using frequencies and percentages, Chi-square or Fisher’s exact test were employed to make comparison. For quantitative variables be summarized with descriptive statistics, the following were presented: N, mean, standard deviation (SD), t-test was employed to compare group and the ANOVA test was allowed a comparison of more than two groups.

The Kaplan-Meier methodology will be used to summarize time-to-event variables. Plots of Kaplan-Meier product limit estimates of time-to-event will be drawn, medians will be presented in addition to confidence intervals, set at 95 percent.

To compare curves for two groups, the log-rank test will be employed.

The level of statistical significance was 5% for all statistical tests (exploratory tests).

To analyzed predictive factor of assessment, logistic regression was used in order to calculate the odds ratio, present with a confidence interval set at 95 percent.

All statistical analyses were conducted with the SAS® System, Version 9.3 (SAS Institute Inc. Cary, NC, USA).

## RESULTS

Between 1/30/21 and 3/26/2021, the application was used by 6 755 unique individuals who completed the baseline questionnaires. Among them, 548 had inclusion criteria and were assessable for outcomes assessment. Figure 1

**Figure 1.**
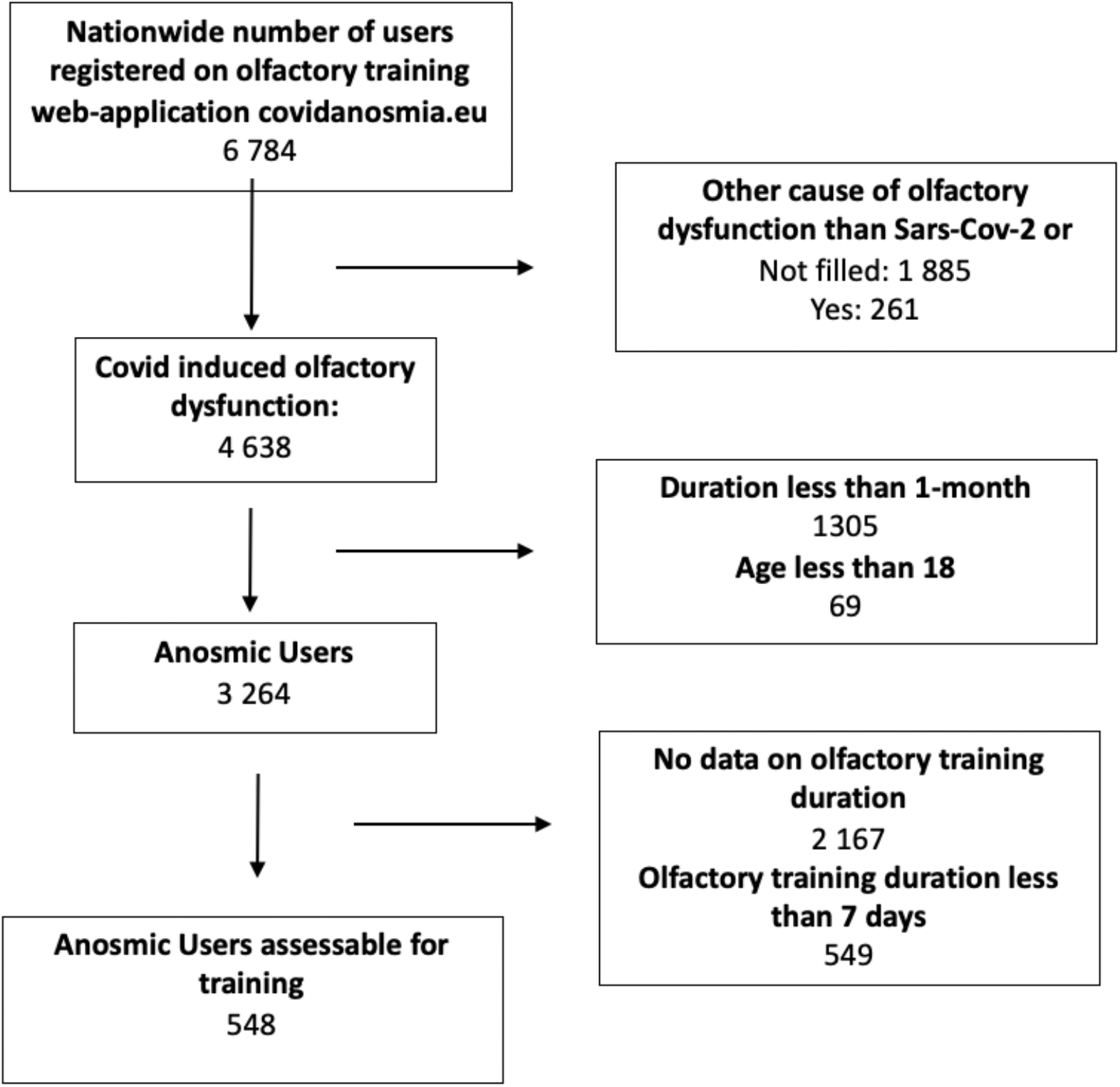
Flowchart of patients using web-application for olfactory training.

Among 548 assessable patients, median age was 42 (min 18, max 84), 65.5% were female, 32.2% estimated having a smell sense more developed than average before olfactory dysfunction, 69.3% estimated than smell sense had an important role in their life and 30.7% did not take care about their smell sense before SARS-CoV-2 infection. Among assessable patients, 20.3% had anosmia (level 0 /10 on olfactory scale), 52.4% had severe hyposmia (level 1 or 2/10 on olfactory scale), 22.8% had moderate hyposmia (level 3-5/10 on olfactory scale), 4.6% had mild hyposmia (level 6-7/10), 50.9% reported a reduction or loss of taste and 52.7% of patients reported parosmia. Patients baseline characteristics are on Table 1. The mean baseline olfactory function of users registered on the web-application with less than 7 days olfactory training or without their last olfactory function assessment on web-application diary was higher than in the studied population (2.23 in 2824 patients and 1.9 in 548 patients, respectively (p Student test <.001).

**Table 1.** Patients characteristics

| Variables | N(%) |
| --- | --- |
| **Sex:** |  |
| Male | 359 (65.5) |
| Female | 189 (34.5) |
| **Smell level before smell dysfunction:** |  |
| In the standard | 354 (64.6) |
| Less developed than average | 18 (3.3) |
| More developed than average | 176 (32.1) |
| **Smell role before lost:** |  |
| Not care about smell | 168 (30.7) |
| Important role | 380 (69.3) |
| **Smell level at baseline:** |  |
| 0 | 111 (20.3) |
| [1-2] | 287 (52.4) |
| [3-5] | 125 (22.8) |
| [6-7] | 25 (4.6) |
| **Olfactory dysfunction duration** |  |
| 1-2 months | 61 (11.1) |
| 2,1 to 3 months | 250 (45.6) |
| 3,1 to 6 months | 167 (30.5) |
| 6 to 12 months | 70 (12.8) |
| **Taste dysfunction:** |  |
| None | 269 (49.1) |
| Dysfunction | 279 (50.9) |
| **Parosmia:** |  |
| No | 259 (47.3) |
| yes | 289 (52.7) |

The mean baseline self-assessed olfactory score was 1.9/10 (SD 1.7) and increased to 4.6 (SD 2.8) after a mean time of olfactory training of 27.7 days (SD 17.2, min 7 days, max 65 days). Olfactory training was associated with at least 1-point increase on olfactory scale in 82.1% of patients (n=450), at least 2-points increase (which was the primary outcome) in 64.2% (n=352) and 49.3% (n=270) at least 3-points increase during the study period. The rate olfactory improvement in patients with olfactory dysfunction anteriority of 12 months was 58.3%. The mean increase in patients having at least 2 points improvement on olfactory scale was 4.1/10 points (SD 1.9).

The duration of the training was associated with better outcome and the kinetic of olfactory function improvement was longer in anosmic patient (olfactory training duration to have 50% probability of improvement = 41 days [36-53]) than in hyposmic patient (33 days [28-36]); p(log-rank) = .0008. Figure 2. There were no differences between severe, moderate and mild hyposmic patients.

**Figure 2.**
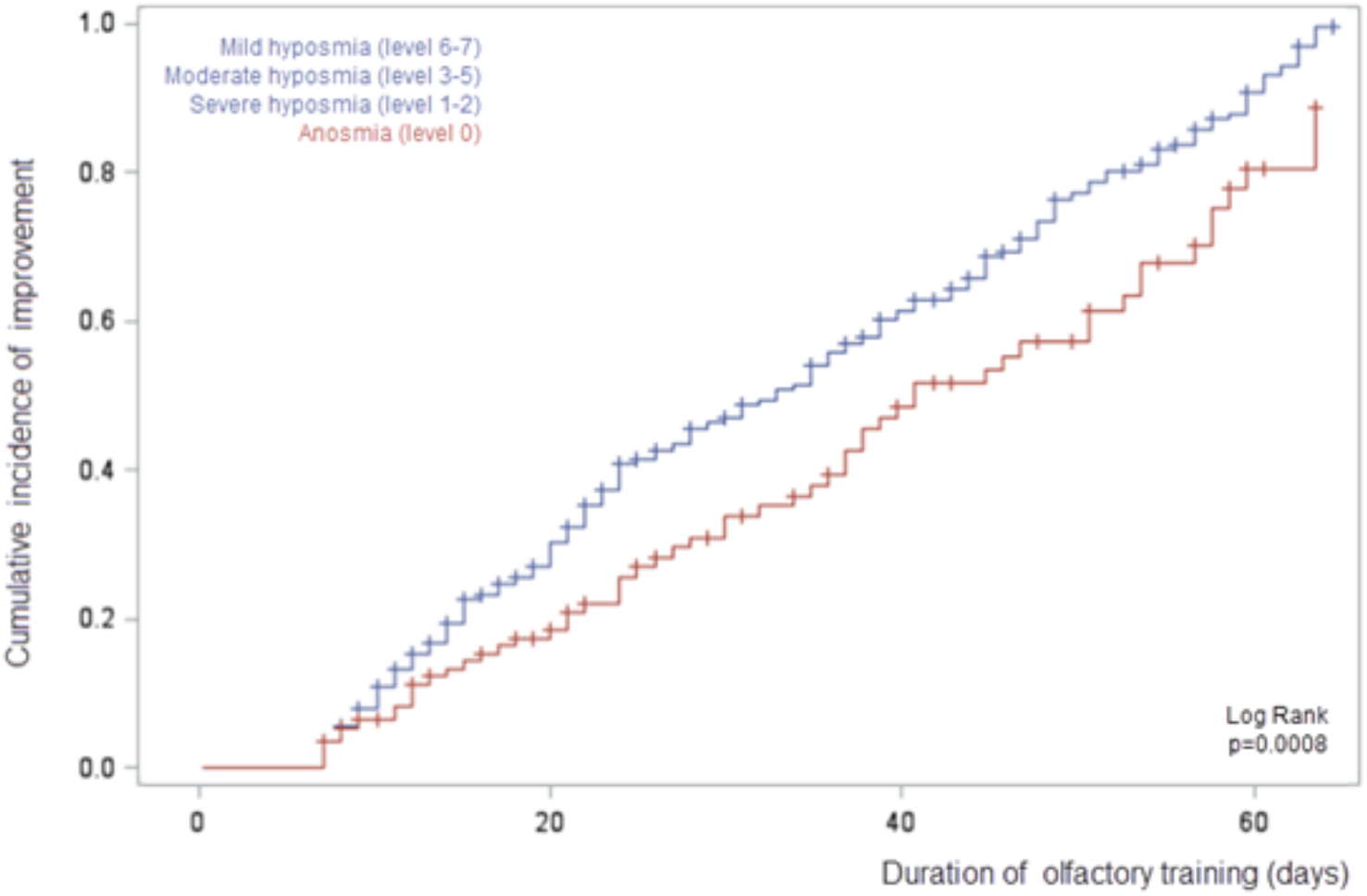
Cumulative incidence of olfactory improvement according to olfactory training duration between anosmic and hyposmic patients. Data of mild, moderate and severe hyposmic patients were pooled in blue curve.

The rate of patients with improvement of at least 2/10 points on olfactory scale was higher in patients training more than 28 days versus patients training less than 28 days (72.2% vs 59.0% respectively, p=.002).

Among patients with 28 days olfactory training or more and who have benefited from an improvement, the mean improvement was 4.4 (SD 2.0) on olfactory scale versus 3.8 points (SD 1.8) in patients with less than 28 days olfactory training (p Student test= .01).

The mean improvement of self-assessed olfactory scale was similar whatever the anteriority of the olfactory dysfunction (p=0.7). Figure 3.

**Figure 3.**
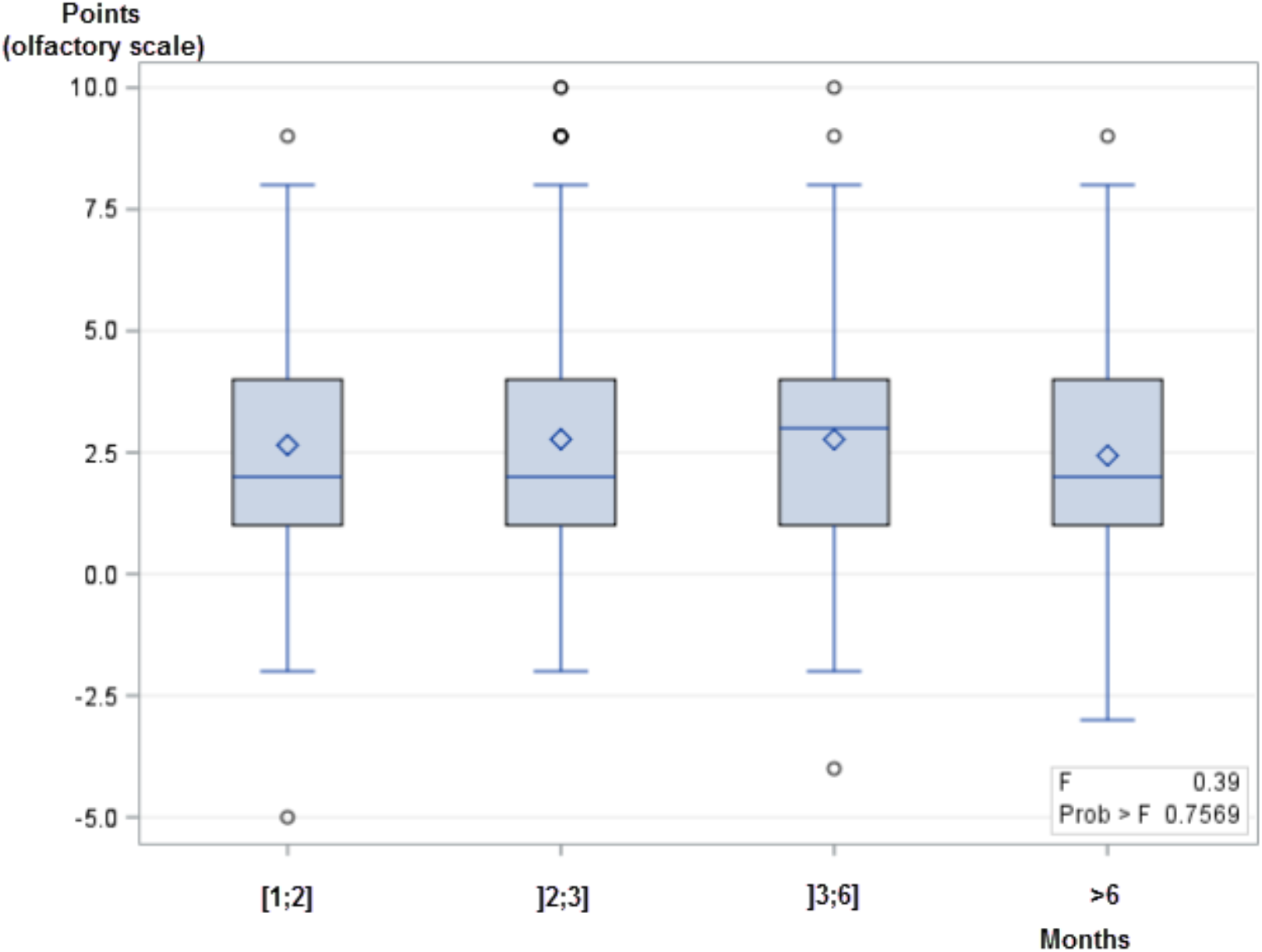
Mean improvement of olfactory function assessed by 0-10 self-assessed olfactory scale after olfactory training according to the duration of persistent olfactory dysfunction.

No predictive factors have been highlighted. Table 2.

**Table 2.** Logistic regression: Assessment of 2 points or more, predictive factors research

|  | Univariate analysis |  |
| --- | --- | --- |
| Variables | RR [95%CI] | p-value |
| **Gender** |  | .7650 |
| Male | 1 |  |
| Female | 0.945 [0.654;1.366] |  |
| Age | 1.012 [0.997; 1.026] | .1081 |
| **Covid tests** |  | .9959 |
| Postive tests | 1 |  |
| No test | 0.934 [0.220; 3.955] |  |
| Yes, but Negative test | 1.058 [0.462;2.425] |  |
| Yes, but no test | 1.074 [0.521; 2.212] |  |
| **Anosmia duration** |  | .8530 |
| More 6 months | 1 |  |
| 1 month | 1.287 [0.627; 2.643] |  |
| 2 months | 1.078 [0.625; 1.861] |  |
| 3-6 month | 1.212 [0.680; 2.160] |  |
| **Smell importance** |  | .6544 |
| In the standard | 1 |  |
| Less than standard | 0.666 [0.256; 1.730] |  |
| Better than standard | 0.909 [0.624;1.325] |  |
| **Taste lost** |  | .4995 |
| No | 1 |  |
| Yes | 1.128 [0.795 ;1.600] |  |
| **Parosmia** |  | .4360 |
| No | 1 |  |
| Yes | 1.149 [0.810 ;1.630] |  |
| **Training duration** |  | .002 |
| <28 days | 1 |  |
| >=28 days | 1.802 [1.247; 2.604] |  |

After a mean time of olfactory training of 28 days, we observed 17.1% of patients (94/548) having an olfactory score superior or equal to 8/10. This high recovery was observed in patients whatever the anteriority of the olfactory dysfunction (p=0.93): 43.6% of these patients had a more than 3-months olfactory dysfunction versus 56.4% who have less than 3-months dysfunction. However, patients with an olfactory training score greater or equal 8, had a baseline score significantly higher than users who did not achieve this level after training. The mean baseline score was 2.9 (SD 2.0) for patients who have high recovery versus 1.6 (SD 1.5) for those who have lower (p<.001).

## DISCUSSION

This study is the first assessing prospectively in real life the benefit of an olfactory training in patients with persistent olfactory dysfunction after SARS-CoV-2 infection. The mean duration of training was 28 days. In this cohort of 548 patients performing the olfactory training assisted by the web-application, an improvement of 2/10 points or more in subjective self-assessed olfactory scale was reported in 64.2% of patients. Beyond 28 days of training, rate of improvement was significantly higher than below this duration, with 72.2% versus 59.0% respectively. The kinetic of improvement was 8-days longer in anosmic patients than in hyposmic. Improvement was observed whatever the anteriority of olfactory dysfunction. A high recovery, that is normal/subnormal self-assessed olfactory function was observed in 16.9% of patients whatever the anteriority of the olfactory dysfunction, but it was more frequent in patients with higher baseline olfactory score (mean score = 2.9/10).

These data are in line with previous randomized report in postinfectious olfactory loss [8]. In this trial, after 18-weeks olfactory training, olfactory function improved in 63% of patients having a duration of olfactory dysfunction of less than 12 months using high concentration oils versus 19% in the control group using low concentration oils. We used high concentration oils combination with the same four odorants used by Damm et al. in two steps for each oil: the first is a blind olfactory stimulation with a given oil like Damm et al. trial; the second, following the first one and with the same oil, is enriched by a visual stimulation consisting of a picture of oil on the smartphone screen delivered by the web-application. We chose this new approach to reinforce the olfactory trial by a mixt olfactory-visual trial because some previous data suggested than human olfactory perception showed substantial benefits from visual cues, suggesting important crossmodal integration between olfactory and visual modalities. [9,12,13] A 4-arms randomized trial assessing the best modalities of training is ongoing to improve olfactory training results [14]. The use of a web-application is a promising tool to improve olfactory training because it allows to associate visual stimulation, video tutorials, provide encouragements and monitor results. It has been shown to be interesting during Sars-Cov-2 pandemic to perform triage of patients and to assess trends of the outbreak at a large scale. [15-17]

Our patients had a persistent anosmia from 2 to 12 months and the rapid benefit observed whatever the anteriority of the anosmia suggests a direct effect of the training. Postinfectious olfactory not caused by SARS-CoV-2 dysfunction is associated with moderate rates of spontaneous recovery. Hendriks reported that spontaneous recovery occurs in 35% of the patients over a period of approximately 12 months. [18] In a retrospective series in 262 subjects with a mean follow up time of 14 months, Reden et al. reported that 32% of olfactory function improvement, assessed with the objective “Sniffin’ Sticks” test, and an increase of at least 6 points in the ‘‘threshold discrimination identification score’’ (TDI score). [19] In a different study, Reden et al. reported clinically relevant improvement of olfactory function in 21% of the participants over a period of approximately 7 months. Hummel et al [7] reported a short-term recovery rate of 6-8% within 4 months, using the same olfactory tests and definitions of improvement as Reden et al. [20] More recently, Havervall et al reported that the duration of olfactory dysfunction after mild SARS-CoV-2 among seropositive health care workers was 14.6%, 10.8 and 9.0% 2, 4 and 8 months after infection, meaning that spontaneous recovery is low. [21]

Spontaneous recovery after persistent olfactory dysfunction in SARS-CoV-2 patients is not well described. Vaira and al reported a mean of 1/10 point on the analogue subjective olfactory scale we used between 30 and 60 days in 138 patients without olfactory training and 20% patients presenting olfactory improvement. Our data suggest that improvement can be obtained tardily after 2-months training. In our study, olfactory training and visual stimulation allowed by a dedicated web-application was associated with 72.2% of patients with 2 points or more improvement after at least 28 days olfactory training and 4.4/10 points mean improvement. [4] In another study, Lechien et al reported that 15.3% and 4.7% of anosmic/hyposmic patients did not objectively recover olfaction at 60 days and 6 months, respectively. The comparison of our study with other studies is however limited because different olfactory tests and scale evaluations were used. [20]

Our study had several limits. There was a lot of excluded patients. We could think that there is a selection bias because patient who don’t feels improvement will more easily stop the training. It could be a confusion factor about the benefit statistically better if patients follow the training more than 28 days. The mean baseline olfactory function of users registered on the web-application with less than 7 days olfactory training or without their last olfactory function assessment on web-application diary was higher than in the studied population (2.23 in 2824 patients and 1.9 in 548 patients, respectively (p Student test <.001). The distribution of the severity of patients suggests a higher severity of patient’s olfactory dysfunction in the population retained in the analysis. This data suggests that the results of olfactory training could be higher in the whole population than in the studied population.

There was no control group, therefore it remains unclear to which percentage spontaneous recovery distorts the results. The scale used to measure olfactory dysfunction and changes was subjective by using a self-assessment analogue scale, they were self-reported and data about olfactory assessment were not confirmed by physician and objective tests. However, the possibility to run olfactory training at home increased the number of recruited patients and triggered high levels of olfactory function recovery compared to spontaneous improvement. Olfactory training associated with visual stimulation assisted by a dedicated web-application is associated with significant olfactive improvement in persistent olfactory dysfunction following SARS-CoV-2 infection, especially if training duration is superior to 28 days.

## Data Availability

Data are not available according to industrial protection.

## Acknowledgment section

We thank users for their participation in this study, as well as Magali Balavoine MSc (Weprom, Angers, France) and the anosmie.org patient’s association.

No one received compensation for their contribution.

## Author Contributions

Pr Denis had full access to all of the data in the study and takes responsibility for the integrity of the data and the accuracy of the data analysis.

## Concept and design

Denis.

## Acquisition, analysis, or interpretation of data

All authors

## Drafting of the manuscript

All authors

## Critical revision of the manuscript for important intellectual content

All authors

## Statistical analysis

Denis, Septans

## Obtained funding

None.

## Administrative, technical, or material support

Denis, Weprom, Kelindi.

## Supervision

Denis, Morinière, Gurden

